# The Role of White Matter Integrity and Neuroplasticity in Stroke Recovery: Insights from DTI and VBM

**DOI:** 10.1101/2025.01.07.25320143

**Authors:** Rongjun Zhang, Zhigang Gong, Wenbing Jiang, Zhaofeng Su

**Author notes:** Correspondence (Rongjun Zhang). These authors contributed equally (Rongjun Zhang, Zhigang Gong).

## Abstract

**Background:** Stroke is a leading cause of disability, significantly affecting the brain’s white and gray matter. Advanced neuroimaging techniques like Diffusion Tensor Imaging (DTI) and Voxel-Based Morphometry (VBM) offer valuable insights into these structural changes.

**Methods:** This study used Tract-Based Spatial Statistics (TBSS) to evaluate white matter integrity in stroke patients using DTI metrics, including Fractional Anisotropy (FA) and Mean Diffusivity (MD). VBM was employed to assess gray matter volume and cortical thickness. Correlation analyses were performed between imaging metrics and clinical scores, such as the NIH Stroke Scale (NIHSS) and Brunnstrom scores.

**Results:** TBSS analysis showed significant reductions in FA (globus pallidus: t = −4.71, p < 0.001; caudate nucleus: t = −4.20, p < 0.001) and increases in MD (globus pallidus: t = 3.96, p < 0.001; caudate nucleus: t = 3.85, p < 0.001) in stroke patients compared to controls. These changes correlated significantly with clinical outcomes; higher FA and lower MD were linked to better motor function (Brunnstrom score: r = 0.90, p < 0.001 for FA in globus pallidus) and lower stroke severity (NIHSS score: r = −0.91, p < 0.001 for FA in globus pallidus). VBM analysis revealed significant gray matter volume increases in the anterior cingulate cortex (t = 4.71, p < 0.05, FWE-corrected) and six other regions at p < 0.001.

**Conclusion:** The study underscores the importance of white matter integrity in post-stroke recovery and highlights neuroplasticity in specific brain regions. Advanced neuroimaging metrics like FA, MD, and gray matter volume are crucial for assessing stroke severity and guiding rehabilitation strategies.

## Introduction

Stroke is a leading cause of global disability, primarily due to damage to the brain’s white and gray matter [1]. Assessing this damage is crucial for understanding stroke pathophysiology and guiding therapy. Advanced neuroimaging techniques, such as Diffusion Tensor Imaging (DTI) and Voxel-Based Morphometry (VBM), offer significant insights into brain structural changes post-stroke [2], providing potential biomarkers for severity and recovery.

DTI quantifies white matter integrity by measuring water molecule diffusion along axonal fibers [3,4]. Key DTI metrics like Fractional Anisotropy (FA) and Mean Diffusivity (MD) reflect white matter tract organization and density [5,6]. Tract-Based Spatial Statistics (TBSS) evaluates these metrics across subjects, minimizing anatomical variability and identifying compromised white matter regions [7].

VBM allows voxel-wise comparison of gray matter (GM) and white matter (WM) volumes across populations [8,9]. It involves segmentation, normalization, and statistical analysis of high-resolution T1-weighted images to detect volumetric differences related to neurological conditions, including stroke [8]. Comparing these volumes between stroke patients and healthy controls highlights stroke-associated structural changes.

Understanding the correlation between structural changes and clinical outcomes is vital for predictive models and rehabilitation strategies. Metrics like FA, MD, and brain volumes are correlated with clinical scores such as the NIH Stroke Scale (NIHSS) [10] and Brunnstrom score [11].

This study used TBSS to assess white matter integrity and VBM to evaluate cortical thickness, GM, and WM volumes in stroke patients versus healthy controls. We aimed to identify significant metric differences and explore correlations between imaging findings and clinical scores. Combining these techniques offers a comprehensive understanding of post-stroke brain structural changes and their clinical implications.

## Methods

### Subject information

This study was approved by the ethics committee of Suzhou Hospital Affiliated to Nanjing University of Chinese Medicine, with written informed consent obtained from all participants. Sixty patients with first-onset unilateral cerebral infarction or intracerebral hemorrhage in the subcortical area (internal capsule or basal ganglia) were recruited. Inclusion criteria included 1-3 months post-onset, stable condition, and exclusion of cerebrovascular malformations or tumors via cranial CTA/MRI. Ages ranged from 35 to 75 years. Exclusion criteria were: MRI contraindications, unstable vital signs, organ failure, or severe cognitive impairment. Additionally, 40 age-matched healthy subjects were enrolled, aged 35-75 years, with no abnormal lesions on routine cranial MRI and no significant cognitive impairment. Exclusion criteria for healthy subjects mirrored those of the stroke group.

### Image Acquisition

All MRI data were acquired with the Philips 3.0 Tesla Ingenia scanner using a standard 8-channel head coil. DTI images were collected using a single-shot echo-planar imaging sequence, with parameters: TR = 4598.5 ms, TE = 82.6ms, 15 non-linear diffusion directions with b = 800 s/mm^2^ and 1 volume with b = 0 s/mm^2^, slice thickness = 2.0 mm, FOV = 224mm × 224mm × 386mm. The 3D T1-weighted structural images were obtained with following parameters: TR = 8.9 ms, TE = 4.1 ms, flip angle = 8°, 260 sagittal slices, slice thickness = 1.0 mm, FOV = 230mm × 230mm × 270mm).

### Tract-Based Spatial Statistics

Tract-Based Spatial Statistics (TBSS) was utilized to analyze white matter integrity in this study. FSL (FMRIB Software Library) was utilized for TBSS. The DTI data underwent preprocessing, which included eddy current and motion correction using FSL’s Diffusion Toolbox (FDT), followed by brain extraction using the Brain Extraction Tool (BET) to remove non-brain tissue. Diffusion tensors were then estimated for each voxel, generating Fractional Anisotropy (FA) and Mean Diffusivity (MD) maps. For the TBSS analysis, all participants’ FA maps were non-linearly registered to the FMRIB58_FA template in standard space. A mean FA image was computed and thinned to create a mean FA skeleton, which represents the central core of the white matter tracts shared across subjects. The FA data from each participant were then projected onto this skeleton, ensuring that the analysis focused on the most consistent and reliable portions of the white matter tracts.

### Voxel-based Morphometry

The VBM analysis was conducted using the Statistical Parametric Mapping (SPM12) software running on MATLAB. The process began with the segmentation of high-resolution T1-weighted images into gray matter (GM), white matter (WM), and cerebrospinal fluid (CSF). These segmented images were then normalized to the Montreal Neurological Institute (MNI) space, a standard anatomical template that facilitates comparison across subjects. During this normalization process, non-linear registration was applied to ensure that individual anatomical differences were minimized, allowing for more accurate group comparisons.

Following normalization, the gray matter and white matter segments were modulated to account for volume changes introduced by the normalization process. This modulation step preserves the original volume information by multiplying the voxel values by the Jacobian determinants derived from the normalization, enabling the detection of regional volume differences rather than just density differences.

The modulated images were then smoothed using an 8-mm full-width at half-maximum (FWHM) Gaussian kernel to enhance the signal-to-noise ratio and to accommodate inter-individual anatomical variability. This smoothing process also ensures that the data conform to the assumptions of the general linear model used in the subsequent statistical analysis.

### Statistical Analysis

Statistical analyses were conducted using IBM SPSS Statistics (version 26.0, IBM Corp.). Pearson’s correlation analysis was utilized to explore the relationships between NIHSS score, Brunnstrom score, cortical thickness, FA, and MD values. Independent samples t-tests were conducted to examine differences in cortical thickness between the patient group and healthy controls. ANOVA analyses were conducted to assess the differences in cortical thickness across ICH, CI, and healthy controls. A p-value of less than 0.05 (two-sided) was considered statistically significant.

## Results

### Demographic characteristics

Upon reviewing the MRI data, imaging quality issues were found in the datasets of some test subjects among the 100 test subjects, making them unsuitable for further analysis. Consequently, the study proceeded with a refined sample. This sample comprised 45 patients, including 29 patients with intracerebral hemorrhage (ICH) and 16 patients with cerebral infarction (CI). Additionally, there were 39 healthy controls (HCs). The average age of the patients was 60.64 years, which was comparable to that of the HCs (57.90 years; t = 1.13, p = 0.2618).

### Identification of FA and MD Changes

Tract-Based Spatial Statistics (TBSS) analysis was employed to identify regions of altered white matter integrity in stroke patients compared to healthy controls. The analysis revealed significant changes in Fractional Anisotropy (FA) and Mean Diffusivity (MD) within specific brain regions. The globus pallidus (GP) and caudate nucleus (CN) were identified as key regions where FA and MD values (Figure 1) were significantly different between stroke patients and controls, suggesting that these areas are particularly vulnerable to white matter damage in stroke.

**Fig 1.**
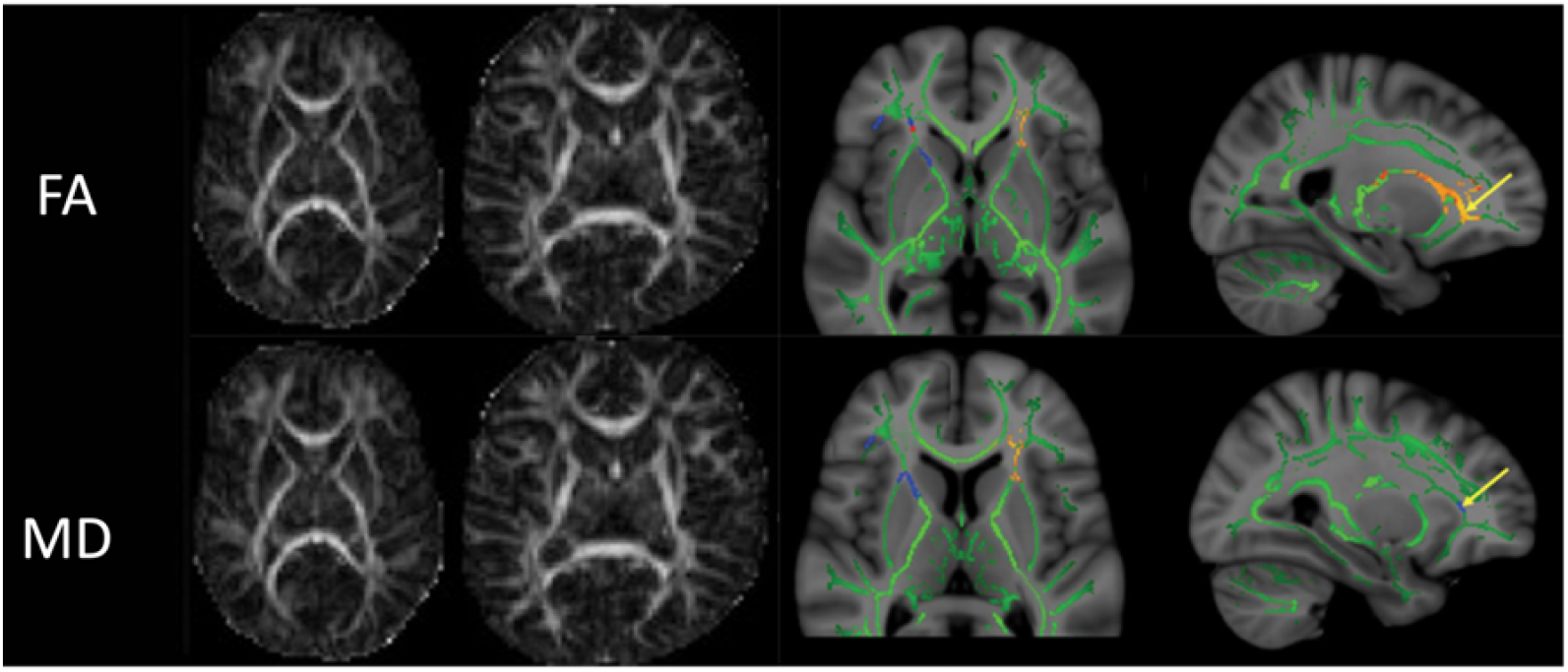
Comparison of Fractional Anisotropy (FA) and Mean Diffusivity (MD) between Stroke Patients and Healthy Controls. The left panel shows axial slices of FA (top row) and MD (bottom row) maps from representative stroke patients and healthy controls. The right panel displays the results of Tract-Based Spatial Statistics (TBSS) analysis, highlighting areas of significant differences in FA and MD values between the two groups. The green skeleton represents the mean white matter tracts common to all subjects. Regions with significant FA reductions in stroke patients compared to healthy controls are shown in blue, while areas with significant MD increases are shown in red-yellow (p < 0.05, corrected for multiple comparisons).

### VBM results

In voxel-by-voxel analysis, only anterior cingulate cortex showed significant alteration in healthy controls versus patients using FWE with p value < 0.05 in the t test. When the threshold was adjusted to an uncorrected p-value of less than 0.001, additional regions demonstrated significant differences. Specifically, seven brain regions showed higher GM volumes in patients compared to healthy controls. These regions include the Left Superior Frontal Gyrus, Left Middle Frontal Gyrus, Right Postcentral Gyrus, Right Inferior Frontal Gyrus, Left Superior Temporal Gyrus, Right Middle Frontal Gyrus, and Superior Parietal Lobule. Notably, when the contrast was reversed (healthy controls > patients), no significant GM reductions were observed in the patient group compared to the controls. These findings suggest that specific cortical regions may undergo volumetric increases following stroke, potentially reflecting compensatory mechanisms or other pathological processes. The precise locations and MNI coordinates of these regions are detailed in Figure2 and Table 1.

**Table 1.** GM alterations detected by VBM.

| $P$ value | Contrast | Location of the peak values | Cluster size<br>(no. of voxels) | MNI coordinates | | | t value of<br>the peak voxels |
| --- | --- | --- | --- | --- | --- | --- | --- |
|  |  |  |  | X (mm) | Y (mm) | Z (mm) |  |
| $P < 0.05$ corr. | PA<sup>a</sup> > HC<sup>b</sup> | Anterior cingulate cortex | 1500 | 3 | 35 | 27 | 4.71 |
|  | HC > PA | - | - | - | - | - | - |
| $P < 0.0005$ uncorr. | PA > HC | Left Superior Frontal Gyrus | 558 | -29 | 60 | -8 | 4.86 |
|  |  | Left Middle Frontal Gyrus | 279 | -42 | 44 | 12 | 4.2 |
|  |  | Right Postcentral Gyrus | 137 | 44 | -24 | 51 | 3.97 |
|  |  | Right Inferior Frontal Gyrus | 218 | 50 | 8 | 24 | 3.96 |
|  |  | Left Superior Temporal Gyrus | 228 | -66 | -29 | -3 | 3.88 |
|  |  | Right Middle Frontal Gyrus | 322 | 42 | 42 | 12 | 3.85 |
|  |  | Superior parietal lobule | 107 | 33 | -56 | 48 | 3.74 |
|  | HC > PA | - | - | - | - | - | - |

<sup>a</sup> Patient group
<sup>b</sup> Healthy group

**Fig 2.**
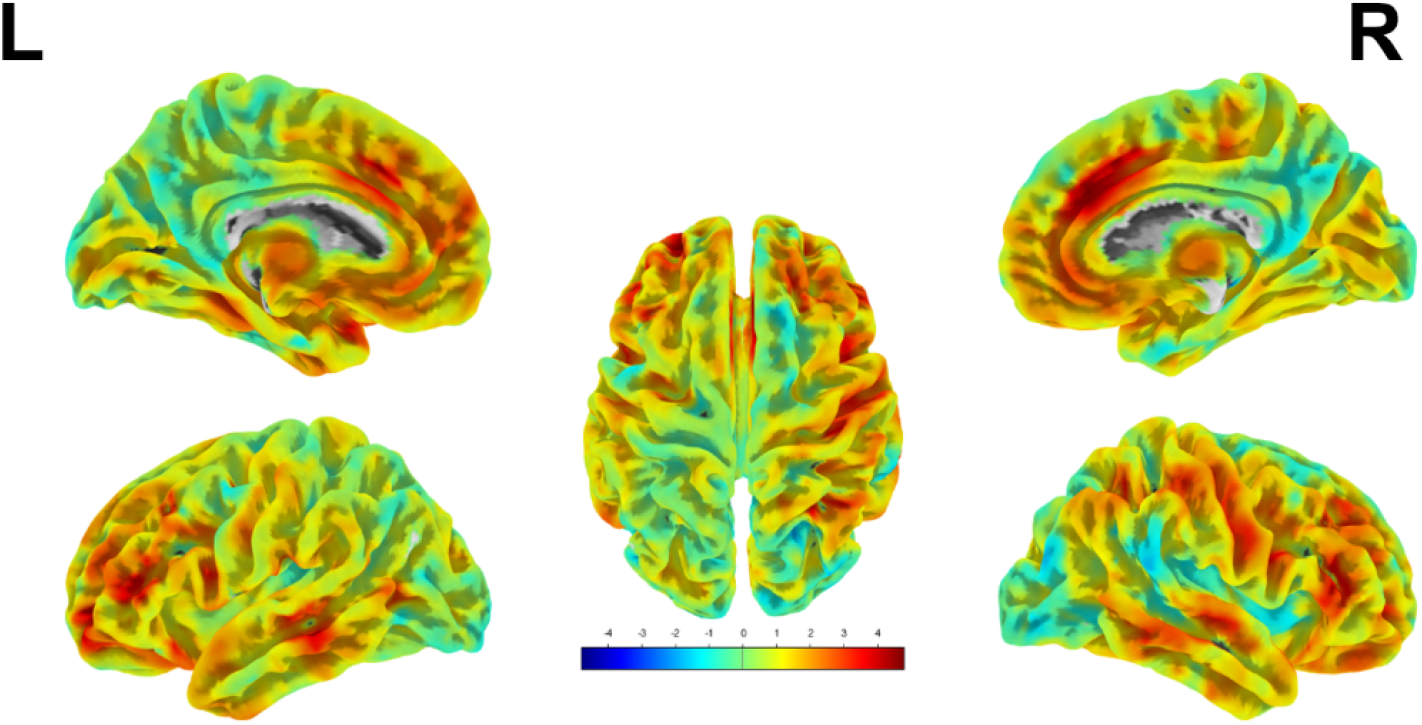
Voxel-Based Morphometry (VBM) Analysis of Gray Matter Volume Differences between Patients group and Healthy Controls. The VBM analysis revealed that, at an uncorrected p-value threshold of 0.001, seven regions exhibited higher GM volume in patients group compared to healthy controls. The color bar represents the t-values, with warmer colors (red-yellow) indicating areas of increased GM volume in patients’ group.

**Fig 3.**
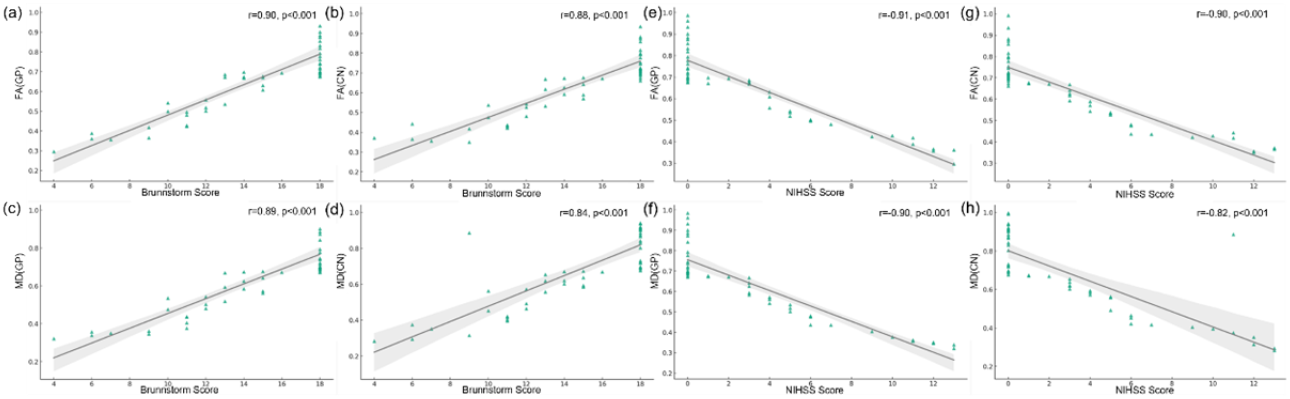
Correlation between Fractional Anisotropy (FA) and Mean Diffusivity (MD) in Specific Brain Regions and Clinical Scores. Scatter plots illustrating the correlations between imaging metrics (FA and MD) in the globus pallidus (GP) and caudate nucleus (CN) and clinical scores (Brunnstrom and NIHSS). (a) and (b) show the positive correlation between Brunnstrom scores and FA in the GP (r = 0.90, p < 0.001) and CN (r = 0.88, p < 0.001), respectively. (c) and (d) depict the positive correlation between Brunnstrom scores and MD in the GP (r = 0.88, p < 0.001) and CN (r = 0.84, p < 0.001), respectively. (e) and (f) show the negative correlation between NIHSS scores and FA in the GP (r = −0.91, p < 0.001) and CN (r = −0.90, p < 0.001), respectively. (g) and (h) illustrate the negative correlation between NIHSS scores and MD in the GP (r = −0.90, p < 0.001) and CN (r = −0.82, p < 0.001), respectively. The shaded regions represent the 95% confidence intervals for the regression lines.

**Fig 4.**
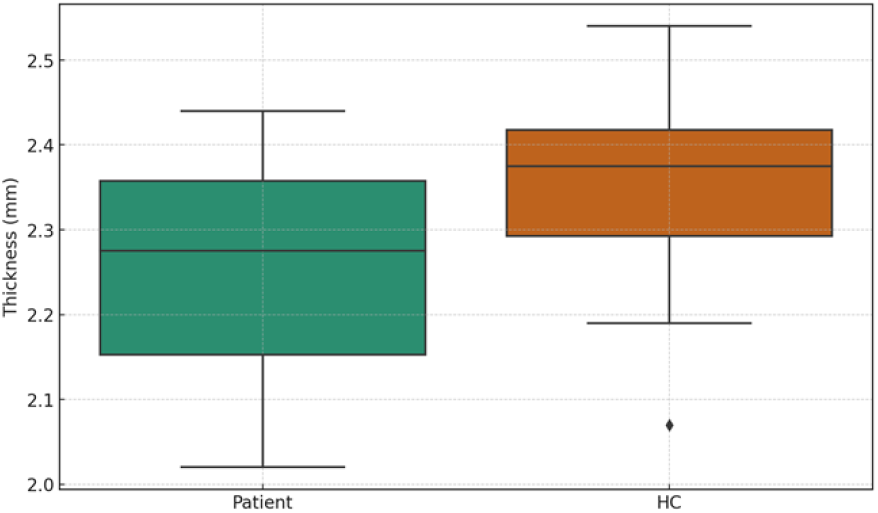
Comparison of Cortical Thickness between Stroke Patients and Healthy Controls. The box plot illustrates the distribution of cortical thickness (in millimeters) for the stroke patient group (left, green) and the healthy control (HC) group (right, orange). The median cortical thickness is represented by the horizontal line within each box, with the boxes showing the interquartile range (IQR) and the whiskers representing 1.5 times the IQR. The plot indicates a statistically significant reduction in cortical thickness in the patient group compared to the healthy controls (p = 0.0056), as determined by an independent samples t-test. The outlier in the HC group is marked by a diamond.

**Fig 5.**
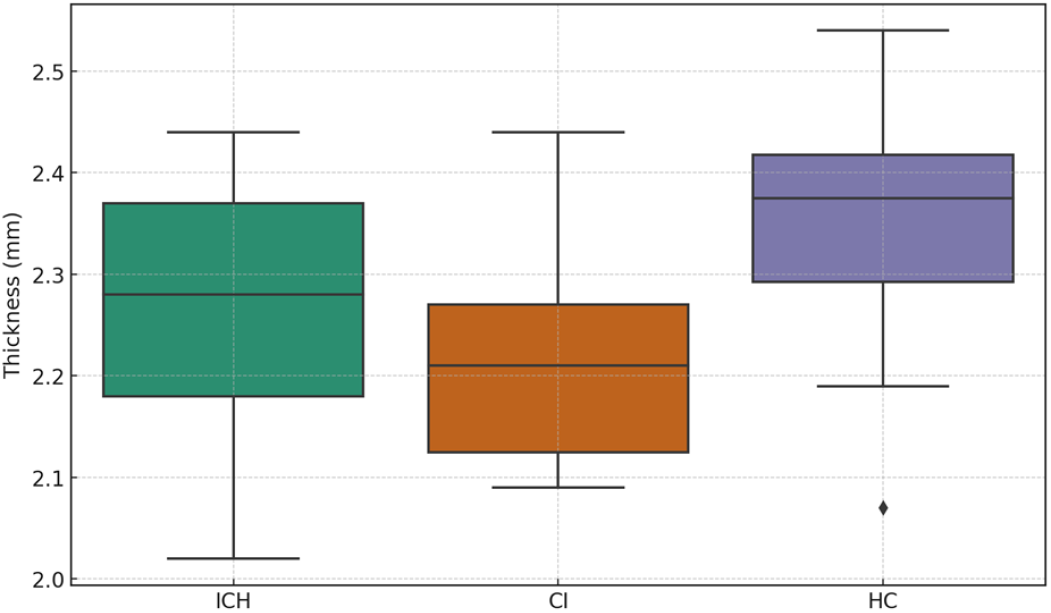
Statistical Parametric Mapping of Cortical Thickness Differences between Stroke Patients and Healthy Controls. The color-coded overlay on the brain surface represents areas with significant cortical thickness differences, with blue indicating regions of decreased thickness and red indicating regions of increased thickness in stroke patients compared to healthy controls. The statistical threshold was set at p < 0.01.

### Correlation Analysis Between Imaging Metrics and Clinical Scores

To explore the clinical significance of the observed white matter changes, correlation analyses were conducted between the FA and MD values in the identified regions (GP and CN) and the clinical scores, including the NIH Stroke Scale (NIHSS) and Brunnstrom scores. The Brunnstrom score showed a strong positive correlation with FA in both the GP (r = 0.90, p < 0.001) and CN (r = 0.88, p < 0.001), as well as with MD in the GP (r = 0.88, p < 0.001) and CN (r = 0.84, p < 0.001), indicating that higher FA and MD values are associated with better motor function. Conversely, NIHSS scores were strongly negatively correlated with FA in the GP (r = −0.91, p < 0.001) and CN (r = −0.90, p < 0.001), and with MD in the GP (r = −0.90, p < 0.001) and CN (r = −0.82, p < 0.001), suggesting that lower FA and MD values correspond to greater stroke severity.

### Cortical Thickness, Grey Matter Volume, and White Matter Volume: Group Comparisons

To evaluate the differences in cortical thickness, grey matter (GM) volume, and white matter (WM) volume between healthy controls (HC) and stroke patients (PA), independent samples t-tests were performed. The results for cortical thickness showed a statistically significant difference between the two groups (t = −2.942, p = 0.0056), indicating that the patients had significantly reduced cortical thickness compared to healthy controls. However, no significant differences were observed in GM volume (t = −1.646, p = 0.107) or WM volume (t = −1.599, p = 0.117) between the two groups, suggesting that stroke did not significantly impact the overall volume of grey or white matter when compared to healthy controls.

Further analysis was conducted using ANOVA to assess differences in cortical thickness, GM volume, and WM volume across three groups: intracerebral hemorrhage (ICH) patients, cerebral infarction (CI) patients, and healthy controls (HC). The analysis revealed significant differences in cortical thickness among the three groups (F = 4.774, p = 0.0132). Post-hoc comparisons indicated that these differences were primarily driven by a reduction in cortical thickness in the stroke groups compared to healthy controls. However, the ANOVA did not reveal significant differences in GM volume (F = 1.290, p = 0.285) or WM volume (F = 1.194, p = 0.312) across the groups, indicating that the type of stroke did not significantly affect these volumetric measures.

## Discussion

In this study, we employed Tract-Based Spatial Statistics (TBSS) and Voxel-Based Morphometry (VBM) to explore the micro and macro structural brain changes associated with stroke. Our findings revealed significant alterations in both white and gray matter structures, which were closely linked to clinical outcomes. The TBSS analysis demonstrated significant reductions in Fractional Anisotropy (FA) and increases in Mean Diffusivity (MD) within the globus pallidus (GP) and caudate nucleus (CN) in stroke patients. These findings are consistent with previous studies that have highlighted the susceptibility of deep gray matter structures to ischemic damage. For instance, studies by Cao et al. (2021) and Xiao et al. (2024) identified the GP and CN as regions with extensive white matter connectivity, making them particularly vulnerable to the disconnection syndrome observed in stroke patients ^[12,13]^. The alterations in FA and MD in these regions likely reflect the breakdown of myelin and axonal integrity, which is critical for maintaining effective neural communication. This aligns with the well-established role of these structures in motor and cognitive functions, both of which are frequently impaired following stroke ^[14,15]^.

Moreover, the strong correlations we observed between FA/MD values and clinical scores further emphasize the clinical relevance of white matter integrity in the GP and CN. Higher FA and lower MD values, which indicate better-preserved microstructural integrity, were positively correlated with higher Brunnstrom scores and negatively correlated with NIH Stroke Scale (NIHSS) scores. These results are consistent with the findings of Yeske et al. (2015), who demonstrated that better white matter integrity in these regions is associated with improved motor recovery and lower stroke severity ^[16]^. The clinical implications of these findings are significant, suggesting that interventions aimed at preserving or restoring white matter integrity in the GP and CN could be crucial for enhancing post-stroke recovery.

The VBM analysis complements our TBSS findings by revealing significant gray matter volume differences between stroke patients and healthy controls. Notably, the anterior cingulate cortex (ACC) showed a significant increase in gray matter volume, even after correcting for multiple comparisons. This finding is intriguing, given the ACC’s well-documented role in cognitive control, emotional regulation, and the processing of complex behaviors ^[17]^. Structural alterations in the ACC have been implicated in a variety of neuropsychiatric disorders, including depression and anxiety, which are common comorbidities in stroke patients ^[18]^. The observed volumetric increase in the ACC might reflect a compensatory mechanism aimed at maintaining cognitive and emotional stability following stroke, a hypothesis that warrants further investigation.

In addition to the ACC, we identified several other regions—such as the superior frontal gyrus, middle frontal gyrus, postcentral gyrus, and superior parietal lobule—where stroke patients exhibited increased gray matter volumes compared to healthy controls. These regions are integral to motor control and sensory processing, which are often disrupted following a stroke ^[19]^. The observed volumetric increases in these regions may represent neuroplastic changes, where surviving brain tissue undergoes hypertrophy to compensate for the loss of function in damaged areas. This aligns with the concept of diaschisis, where remote brain regions undergo structural and functional changes in response to localized injury ^[20]^.

Interestingly, when we reversed the contrast (healthy controls > stroke patients), no significant reductions in gray matter volume were observed in the stroke patients. This finding challenges the traditional view that stroke primarily results in atrophic changes and suggests instead that the brain may engage in compensatory volumetric increases in certain regions ^[21]^. This neuroplastic response could be a target for therapeutic interventions aimed at enhancing recovery. For example, studies have shown that rehabilitation strategies that stimulate neuroplasticity—such as constraint-induced movement therapy and transcranial magnetic stimulation—can lead to structural brain changes and improve functional outcomes in stroke patients ^[22,23]^. Our findings suggest that such interventions may be particularly effective if they are tailored to target the regions identified in our VBM analysis.

The absence of significant gray matter reductions in stroke patients, despite the presence of functional impairments, highlights the complexity of stroke pathology. It suggests that functional deficits may arise not solely from tissue loss but also from disconnections between brain regions, as well as maladaptive neuroplasticity. This is supported by findings from functional MRI studies, which have shown that stroke can lead to widespread alterations in brain connectivity, even in regions distant from the lesion site ^[23,24]^. These disconnections can disrupt the integration of information across brain networks, leading to deficits in motor control, cognition, and other functions ^[25]^.

The findings from this study have important clinical implications. The strong associations between imaging metrics and clinical outcomes suggest that DTI and VBM could serve as valuable tools for assessing stroke severity and predicting recovery trajectories. By identifying specific brain regions that are most affected by stroke, these imaging techniques could also inform the development of targeted rehabilitation strategies aimed at optimizing recovery. For instance, our findings suggest that interventions that promote white matter integrity in the GP and CN, or that enhance gray matter volume in regions such as the ACC and frontal gyri, could be particularly beneficial for stroke patients.

However, it is important to acknowledge the limitations of this study. The relatively small sample size may limit the generalizability of our findings, and the cross-sectional design precludes the ability to make causal inferences about the observed relationships. Longitudinal studies with larger cohorts are needed to validate our findings and to explore the temporal dynamics of brain changes following stroke. Additionally, integrating these imaging metrics with functional assessments and other biomarkers could provide a more comprehensive understanding of the mechanisms underlying stroke recovery.

In conclusion, this study highlights the critical role of advanced neuroimaging techniques in uncovering the structural brain changes associated with stroke. Our findings underscore the importance of preserving white matter integrity and leveraging neuroplasticity to improve functional outcomes. Future research should focus on longitudinal analyses and the development of targeted interventions that capitalize on the brain’s inherent capacity for adaptation and recovery.

## Data Availability

Data cannot be shared publicly because of privacy. Data are available from the Institutional Data Access / Ethics Committee (contact via corresponding author) for researchers who meet the criteria for access to confidential data. The data underlying the results presented in the study are available from (include the name of the third party and contact information or URL).

